# Decentralized, privacy-preserving surgical video analysis with Swarm Learning

**DOI:** 10.1101/2025.10.02.25337106

**Authors:** Oliver Lester Saldanha, Kevin Pfeiffer, Sebastian Bodenstedt, Max Kirchner, Alexander C. Jenke, Catarina Barata, Silvia Barbosa, Julia Barthel, Matthias Carstens, Laura Teixeira Castro, Karolin Dehlke, Sophia Dietz, Sotirios Emmanouilidis, Guido Fitze, Martin Freitag, Fabian Holderried, Weam Kanjo, Linda Leitermann, Sören Torge Mees, Antonio S. Soares, Margarida Pascoal, Steffen Pistorius, Conrad Prudlo, Jurek Schultz, Astrid Seiberth, Karolin Thiel, Xuewei Wu, Daniel Ziehn, Stefanie Speidel, Jürgen Weitz, Marius Distler, Jakob Nikolas Kather, Fiona R. Kolbinger

**Author notes:** Corresponding author: Fiona R. Kolbinger. Co-senior authors: Jakob Nikolas Kather and Fiona R. Kolbinger.

## Abstract

**Background:** Progress in artificial intelligence-based analysis of surgical videos has been constrained by reliance on manual frame-level annotations rather than patient-level outcomes. In addition, concerns about data privacy restrict the exchange of laparoscopic video data and, thereby, multicenter collaboration.

**Methods:** To address these limitations, we developed a pipeline that integrates weakly supervised deep learning with Swarm Learning, a decentralized machine learning approach that enables collaborative model training without data centralization. We evaluate our pipeline using a newly curated dataset of 397 laparoscopic appendectomy recordings from six international surgical centers. We identified optimal modelling configurations (frame sampling rates and model architectures) and subsequently compared Swarm Learning to single-center and centralized learning across three novel patient-level disease staging tasks: (i) binary detection of perforated appendicitis, (ii) laparoscopic grading of appendicitis, and (iii) histopathologic inflammation grading. In addition, we surveyed participating centers to identify real-world barriers to the clinical implementation of our decentralized learning pipeline for surgical video analysis.

**Results:** For appendicitis grading tasks, frame sampling at 1.0 frames per second and use of the SurgTempoNet architecture resulted in reliable classification performance, outperforming SurgFrameNet and Multiple Instance Learning. Across all three disease staging tasks, Swarm Learning consistently outperformed single-center training and achieved performance comparable to centralized learning, with stable generalization in external validation. The user survey identified hardware failure and limited integration of the decentralized learning pipeline with electronic patient records as key barriers to the clinical implementation of our decentralized learning pipeline for collaborative surgical video analysis.

**Conclusions:** Weakly supervised deep learning enables the prediction of patient-level endpoints directly from surgical video data. Swarm Learning facilitates privacy-preserving multicenter collaboration and achieves performance on par with centralized learning, highlighting its potential for advancing clinically relevant, collaborative AI development in surgical video analysis, especially when integrated with patients’ electronic health records.

**Article Description:** This study introduces a decentralized, privacy-preserving pipeline that combines weakly supervised deep learning with Swarm Learning to predict patient-level outcomes from laparoscopic appendectomy videos. Using data from six international surgical centers, the approach demonstrated performance comparable to centralized learning across three disease staging tasks while preserving data confidentiality by design.

## Introduction

Artificial Intelligence (AI)-based analysis of surgical video data holds potential to augment laparoscopic surgery by automating documentation^1^ and providing quality control, objective feedback, and intraoperative decision support^2–4^. Several such tools have been proposed in recent years, for example, to visualize anatomical risk and target structures during surgeries^5–9^, identify surgical ac-tions^10–12^, or analyze surgeon skill^13–15^. Despite academic progress, current AI-based approaches to surgical video analysis have not advanced beyond early-stage feasibility studies in clinical settings. Moreover, none have demonstrated measurable benefits for clinicians or patients, such as reductions in surgical duration or complication rates^16,17^.

Two factors critically limit progress in AI-based surgical video analysis: Limited availability of data due to inconsistent recording of surgical procedures, which is not part of the clinical routine at most institutions^18^, and a lack of annotations for laparoscopic video data, which are typically added to individual video frames in a time-consuming manual process that requires expert knowledge^19^. As a result, most existing works in AI-based surgical video analysis rely on strongly supervised analysis of manually annotated single-center datasets and lack reproducibility.^16^ Between the few institutions that have access to annotated datasets of surgical videos, there is currently limited collaboration due to data privacy concerns.^20,21^ This consideration is particularly relevant in surgical video analysis, as surgery videos, in contrast to other medical imaging types such as radiological or endoscopic imaging, comprise not only patient-related data but also surgeon-related information, such as tool trajectories, that relate to surgeon skill.

The use of patient-level labels, such as disease stage or patient outcomes, that are readily available from electronic health records in a weakly supervised learning scheme offers two advantages over the current standard practice of frame-level annotations and predictions: First, it drastically reduces the annotation effort while increasing the objectivity and, thus, reliability of labels. Second, the prediction of patient outcomes rather than descriptive video metadata, such as frame-level instrument visibility, increases the clinical value of the resulting models.

Federated learning approaches have been proposed to avoid data centralization in multicenter AI training: Independent models are trained at participating institutions, and model weights are communicated between institutions, resulting in collaboratively trained prediction models without the risk of confidentiality breaches.^22^ However, federated learning requires a central coordinator to consolidate the independently trained models. In healthcare, avoiding a central coordinator enhances privacy and security of patient data, rendering respective methods more suitable than conventional federated learning approaches. In Swarm Learning, a modern decentralized learning method, model parameters are exchanged within the Swarm network using a private, permissioned blockchain based on an open-source implementation of Ethereum^23^. This process is not governed by a central coordinator, resulting in a democratic representation of all participating centers in the collaborative AI training process. Swarm Learning has been evaluated on a range of medical data types, including genomic data^23^, radiological^24^, and histopathological^25^ imaging, but not on dynamic data, such as surgery recordings, that introduce distinct challenges, such as temporal dependencies between frames, varying camera positions and orientations, smoke and fluid interference, and substantial variations in procedure duration.

Using a novel dataset of video recordings and clinical metadata from six international surgical centers, we present and benchmark technical approaches that integrate weakly supervised learning with Swarm Learning to facilitate patient-level predictions from surgical video data, using three novel patient-level disease staging tasks (**Figure 1a, b**) as examples: (i) detection of perforated appendicitis (binary prediction task), (ii) laparoscopic grading of appendicitis, and (iii) histopathologic inflammation grading.

**Figure 1:**
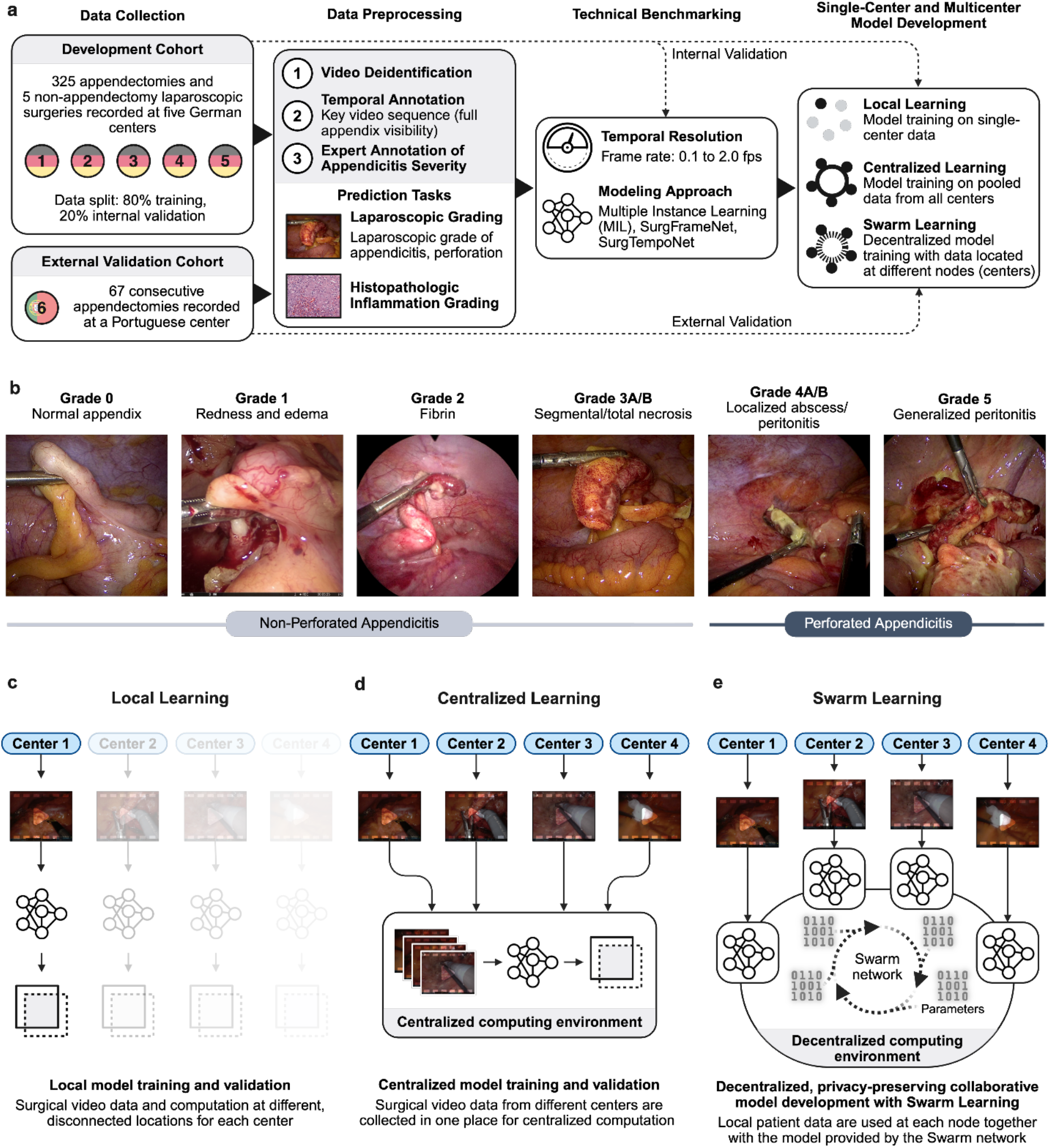
Prediction of patient-level outcomes from surgical video data with Swarm Learning. **(a)** Study overview. Data from six clinical centers were deidentified and labeled with temporal and class labels of appendicitis severity. Our systematic benchmarking effort identified a suitable temporal resolution and modeling approach for subsequent experiments comparing local learning, centralized learning, and Swarm Learning for all three prediction tasks. **(b)** The laparoscopic grade of appendicitis was annotated based on the Gomes classification^26^, as described previously.^27^ **(c)** Local learning, with data and computation at disconnected locations, is common practice in surgical data science, resulting in most studies considering small, single-center cohorts. **(d)** A centralized process collects data and computing processes in one location. **(e)** In Swarm Learning, each center represents a node in the Swarm network, where local patient data are used in conjunction with the model provided by the Swarm network to update the collaboratively trained model without the direct exchange of patient data. In this study, we present a proof-of-concept for the decentralized, collaborative training of prediction models for patient-level outcomes on surgical video data with Swarm Learning, and compare Swarm Learning to local and centralized model training.

Our study reports three key insights: First, to determine optimal modelling parameters for the novel patient-level prediction tasks, we comprehensively benchmark the performance of a range of temporal sampling strategies and weakly supervised image analysis architectures. Second, we compare model performances of local learning, the current standard in AI-based surgical video analysis (**Figure 1c**), centralized learning (**Figure 1d**), and decentralized, privacy-preserving Swarm Learning (**Figure 1e**). Third, through a survey of the participating clinical institutions, we identify real-world challenges to a broad clinical implementation of decentralized AI training on surgical video data.

## Methods

### Ethics statement

We conducted this study according to the Declaration of Helsinki and its later amendments. The responsible Institutional Review Boards reviewed and approved this study on August 4, 2022 (TUD Dresden University of Technology, approval number BO-EK-332072022), September 13, 2023 (Sächsische Landesärztekammer, approval number EK-BR-75/23-1), December 23, 2023 (Landesärztekammer Baden-Württemberg, approval number B-F-2023-023), and November 15, 2023 (Hospital Prof. Doutor Fernando Fonseca, approval number 113/2023). Our study was prospectively registered at the German Clinical Trials Register (Deutsches Register Klinischer Studien, DRKS) on December 9, 2022 (trial registration ID: DRKS00030874). Following local legislature, no written informed consent was required for anonymized data acquisition, data analysis, and publication of results.

### Patient cohorts

A total of 392 patients with suspected acute appendicitis undergoing laparoscopic appendectomy at six independent clinical centers were recruited. In addition, five patients undergoing non-appendectomy laparoscopic surgery were recruited as negative controls, resulting in a total of 397 laparoscopic surgery recordings analyzed in this study. Five German centers provided data for the development cohort used for model implementation and internal validation. One Portuguese center (Hospital Prof. Dr. Fernando Fonseca, Lisbon) provided data for external testing. All included patients had a clinical indication for the surgical procedure. **Table 1** summarizes the patient characteristics and metadata.

**Table 1:**
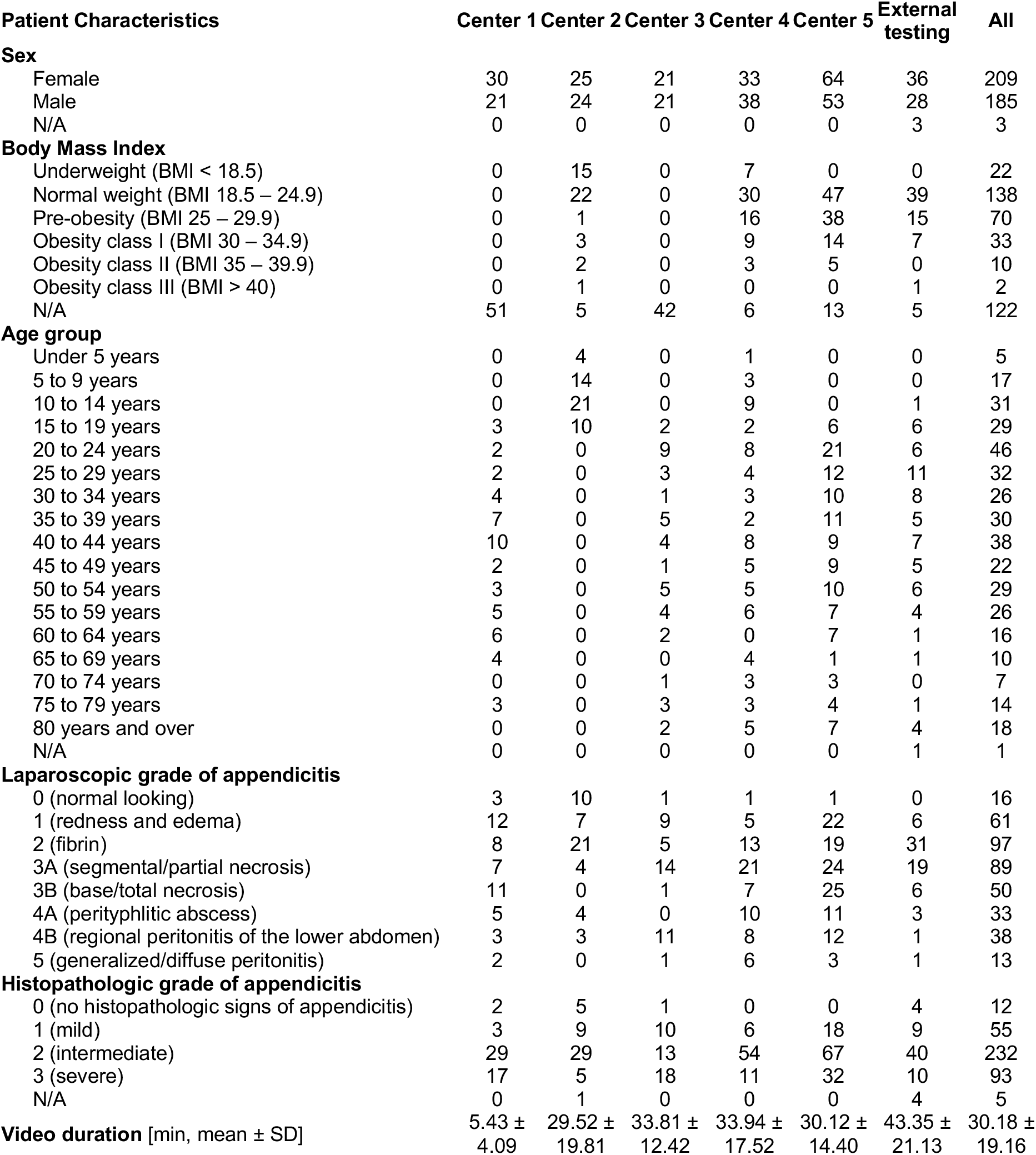
Summary of patient characteristics and metadata. Video durations are provided as mean ± standard deviation. Abbreviation: Body-mass index (BMI).

A detailed data descriptor outlining the recording, preprocessing, annotation, and technical validation process is available at https://doi.org/10.1101/2025.09.05.25335174^27^, summarized in **Supplementary Information 1**.

### Experimental setup

In this work, we present comprehensive evaluations of three patient-level prediction tasks from laparoscopic appendectomy video recordings: (i) detection of perforated appendicitis (binary prediction task), (ii) laparoscopic grading of appendicitis, and (iii) histopathologic inflammation grading. Our experimental analysis comprises two parts: First, a technical benchmarking to determine optimal modelling parameters (**Supplementary Information 2, Supplementary Information 3**), and second, using the best-performing configuration, a comparative analysis of local learning, centralized learning, and decentralized, privacy-preserving Swarm Learning (**Supplementary Information 4**), across all three patient-level prediction tasks.

The experimental pipeline consisted of four distinct phases. First, individual models were trained on each center’s 80% training subset for all three classification tasks. Second, training subsets from all five centers were merged to create unified classifiers trained on the combined dataset. Third, a Swarm Learning approach was implemented using three separate bare-metal servers, with two nodes containing data from two centers each and one node with a single center dataset. Finally, all models underwent validation on both the internal validation set, comprising the combined 20% internal validation data subsets, and on the independent external testing dataset for generalizability assessment. All experiments were repeated three times using specified random seeds.

### Hardware for data acquisition, processing, and training

Local training and Swarm Learning experiments were conducted on a dedicated computing workstation operating Ubuntu 22.04.5 LTS (x86_64 architecture). The system comprised an Intel Xeon W-2245 processor, 128 GB RAM, and an NVIDIA Quadro RTX 6000 graphics processing unit with 24 GB dedicated video memory.

### Statistics

The primary statistical endpoint for classification performance was the AUROC. The AUROCs of three training runs (technical repetitions with different random starting values) of a given model were compared using DeLong’s test with a significance level of p<0.05. AUROCs are reported as mean ± standard deviation. Additionally, we evaluated model performance using the F1 score for the binary classification task.

### Survey to identify practical challenges for clinical adoption of Swarm Learning

To identify key challenges for the clinical adoption of Swarm Learning, participating physicians at the five centers contributing to the development cohort were surveyed between June 14, 2024 and June 15, 2025, following the completion of local data collection. Specifically, centers were asked to rate the technical reliability, practical feasibility, and the potential for errors and data loss related to different system and process components (i.e., patient identification, video recording, data transfer), on a five-point Likert scale. Additionally, center representatives estimated the time investment per patient, as well as the time required for technical troubleshooting, throughout the entire data collection period. Survey results are reported using descriptive statistics and frequency distributions.

### Data availability

A detailed data descriptor outlining the recording, annotation, and technical validation process is available at https://doi.org/10.1101/2025.09.05.25335174^27^. The development cohort multicenter dataset of appendectomy recordings and corresponding clinical metadata will be made publicly available upon acceptance of this work.

### Code availability

All source code is available at https://github.com/KatherLab/swarm-learning-hpe/tree/surgery_swarm. The code is based on and requires the HPE implementation of Swarm Learning, which is publicly available at https://github.com/HewlettPackard/swarm-learning/releases/tag/v2.2.0.

## Results

### Benchmarking of temporal resolution and modeling strategies for the detection of perforated appendicitis

For the binary detection of perforated appendicitis from surgical video data, prediction performance increased at higher frame sampling rates, and the SurgTempoNet models consistently outperformed SurgFrameNet and Multiple Instance Learning models. Among all evaluated models, the SurgTempoNet configuration using 1.0 fps achieved the highest AUROC of [0.918] ± [0.007] (**Figure 2a, Supplementary Information 2, Supplementary Tables 1-3**). Therefore, this configuration was selected for all subsequent experiments.

**Figure 2:**
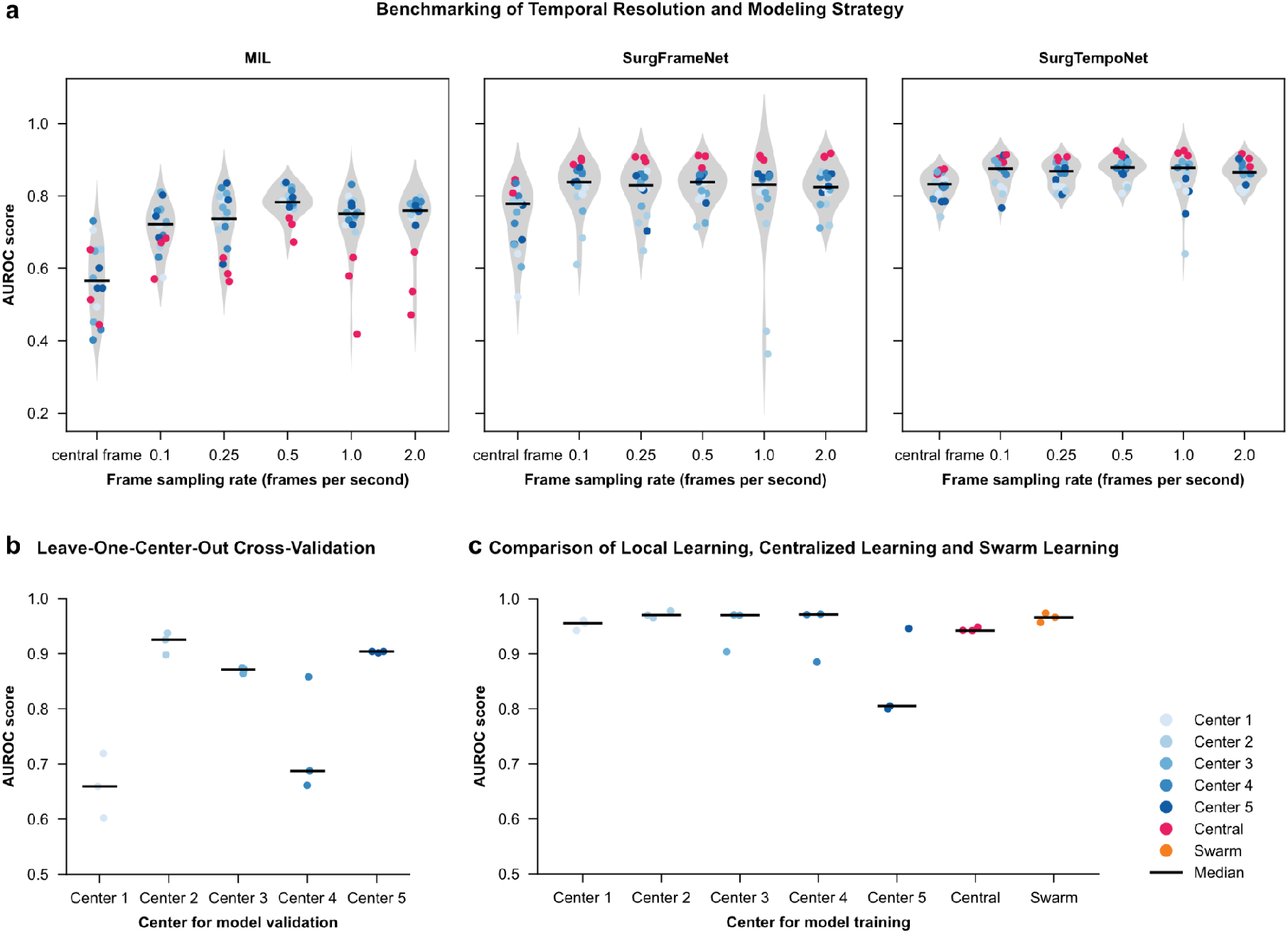
Detection of perforated appendicitis from appendectomy video recordings. The figure illustrates the performance of binary classification models for the detection of perforated appendicitis across internal **(a)**, cross-center **(b)**, and external validation **(c). (a)** Internal validation AUROC scores across five centers and all-center pooled data, comparing Multiple Instance Learning (MIL), SurgFrameNet, and SurgTempoNet models using frames sampled at varying frame rates. **(b)** Leave-one-center-out cross-validation of SurgTempoNet models with input frames sampled at 1.0 frames per second. Each center was held out once for validation while models were trained on data from the remaining centers. **(c)** External validation of SurgTempoNet models trained on data from individual centers, centralized data, and via decentralized Swarm Learning, evaluated on an unseen external dataset. In all plots, dots and the black line indicate individual experiments and the median performance, respectively. Abbreviation: Area under the receiver operating curve (AUROC).

### Evaluation of cross-institutional generalizability of patient-level prediction models for the detection of perforated appendicitis

To evaluate the cross-institutional generalizability of patient-level prediction models for the detection of perforated appendicitis from surgery recordings, we evaluated their model performance in a leave-one-center-out cross-validation scheme. Overall, we observed considerable differences across the five centers, with mean AUROC and F1 scores varying between [0.660] ± [0.058] and [0.474] ± [0.053], respectively (validation on Center 1) and [0.920] ± [0.020] and [0.587] ± [0.070], respectively (validation on Center 2), indicating relevant heterogeneity in laparoscopic procedure recordings between centers and implying limitations in cross-institutional model generalizability. (**Figure 2b**). The high validation performance on data from pediatric patients (Center 2) indicates that differences in patient age distribution do not negatively impact model performance (**Figure 2b, Table 1**).

### Comparative analysis of local learning, centralized learning, and Swarm Learning for the detection of perforated appendicitis

We validated the SurgTempoNet model for detection of perforated appendicitis on an independent external patient cohort and compared models trained locally (i.e., on single-center data), via centralized learning, and via Swarm Learning (**Figure 1a**).

The Swarm Learning-based ensemble consistently matched or exceeded the performance of most individual-center models, achieving an AUROC of [0.966] ± [0.009] and an F1-score of [0.636] ± [0.135]. The centralized model achieved similarly strong performance (AUROC of [0.944] ± [0.004]; F1-score of [0.596] ± [0.047]). Both the centralized and Swarm Learning models demonstrated markedly reduced performance variability on external validation compared to single-center models (**Figure 2c, Supplementary Table 4**). These findings highlight the advantages of Swarm Learning in enhancing model generalizability and robustness across varied clinical settings while retaining data privacy by design.

### Comparative analysis of local learning, centralized learning, and Swarm Learning for laparoscopic appendicitis grading from surgical videos

Across cohorts, the distribution of laparoscopic grades of appendicitis varied strongly (**Figure 3a, Table 1**). The internal validation for multiclass laparoscopic grading revealed variations in model performance by grade (**Supplementary Table 5**). The Swarm Learning model performed well across grades, achieving particularly reliable detection for Grade 1 (AUROC of [0.878] ± [0.006]) and Grade 4 (AUROC of [0.809] ± [0.016]) (**Figure 3b**). Overall, both the Swarm Learning and centralized models outperformed individual-center models for laparoscopic grading of appendicitis in the internal validation at average AUROCs of [0.731] ± [0.013] and [0.756] ± [0.017], respectively (**Figure 3b**).

**Figure 3:**
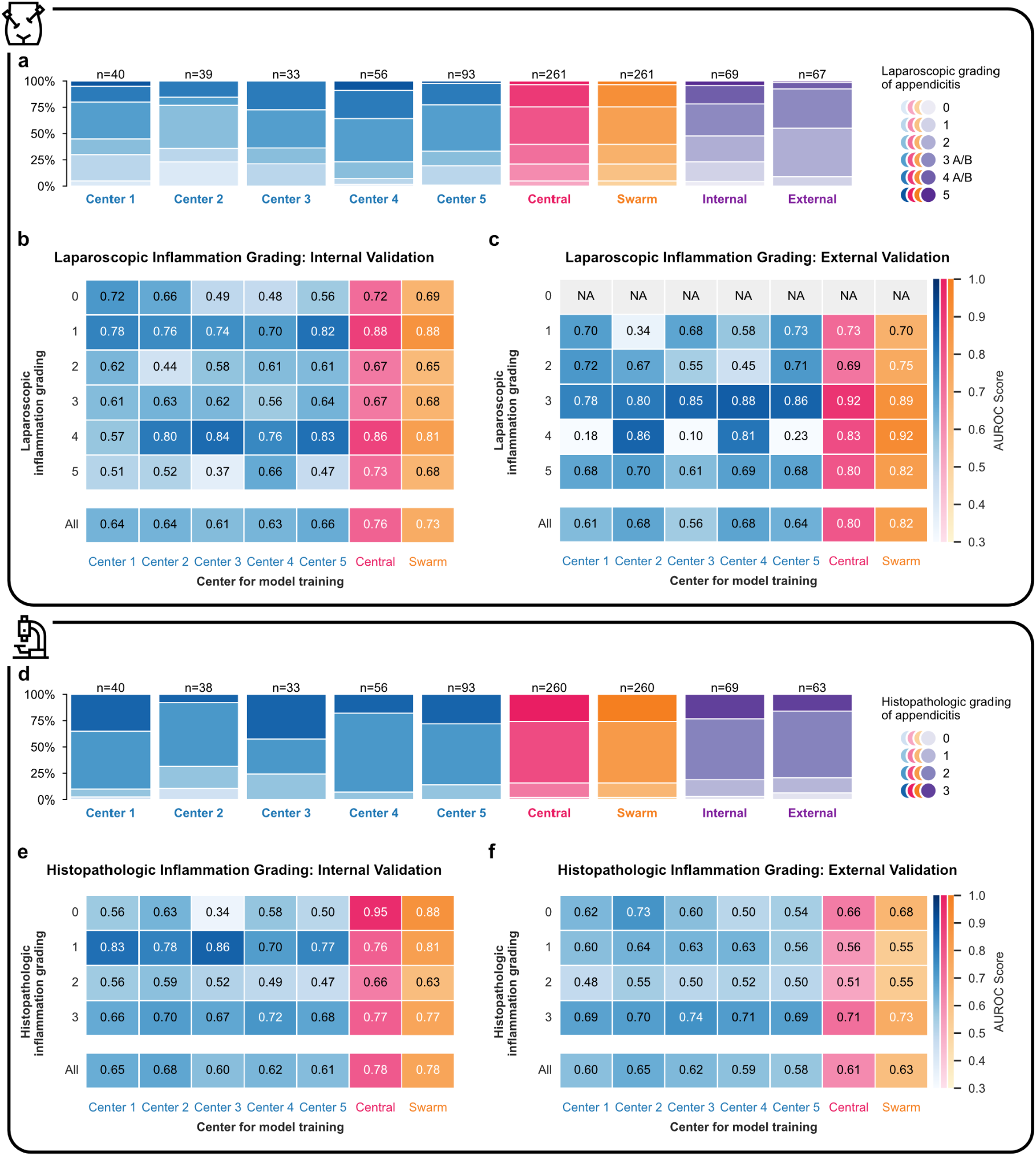
Multicenter model performance for laparoscopic (top panel) and histopathologic (bottom panel) grading of appendicitis. The figure illustrates the data distribution and classification performance (area under the receiver-operator characteristic curve, AUROC) of SurgTempoNet models for laparoscopic grading **(a-c)** and histopathologic inflammation grading **(d-f)** of appendicitis, trained via local learning (single-center data), centralized learning, and Swarm Learning. **(a)** Stacked bar charts displaying class distribution for laparoscopic grading across training subsets of each center in the development cohort (blue), pooled training subsets for centralized (pink) and Swarm Learning (orange) approaches, as well as internal and external testing datasets (purple). Segments represent instances per laparoscopic grade (Grades 0–5). **(b)** Heatmap of internal validation AUROC scores for laparoscopic grading across models trained on single-center data, centralized data, and via Swarm Learning (combined cross-center ensemble). Each cell indicates the AUROC for a specific grade (rows: Grades 0–5) per training setup (columns). The bottom row reports the average AUROC across grades for each training setting. **(c)** Heatmap of external validation AUROC scores for the same laparoscopic grading task. Models trained on single-center data, centralized data, and via Swarm Learning were tested on unseen external data. Axes and colormaps follow the same conventions as panel (b). **(d)** Stacked bar charts illustrating class distributions of histopathologic inflammation grades (Grades 0–3) for each training subsets of each center in the development cohort (blue), pooled training subsets for centralized (pink) and Swarm Learning (orange) approaches, as well as internal and external testing datasets (purple). Segments represent instances per histopathologic grade (Grades 0–3). **(e)** Heatmap showing internal validation AUROC scores for multiclass histopathological staging, using the same structure as panel (b). Rows correspond to four histopathologic inflammation grades (Grades 0–3), columns represent individual and aggregate training setups. **(f)** External validation heatmap for histopathologic inflammation grading. AUROC scores are shown for each stage across models trained at individual centers as well as via centralized and Swarm Learning approaches.

In the external validation, we observed substantial center-wise variation in AUROC performance across Grades 0 to 5 for local models. In contrast, the Swarm Learning model achieved higher and more consistent AUROCs of [0.701] ± [0.015], [0.746] ± [0.008], [0.892] ± [0.020], [0.924] ± [0.033], and [0.825] ± [0.017] for Grades 1 to 5, respectively. These performances were similar to those of the centralized model (**Figure 3c, Supplementary Table 5**). Collectively, these findings indicate that Swarm Learning not only improves overall classification accuracy but also stabilizes performance across challenging grades and heterogeneous institutional datasets, underscoring the advantages of ensemble approaches in complex patient-level grading tasks based on surgical video input.

### Comparative analysis of local learning, centralized learning, and Swarm Learning for the histopathologic appendicitis grading from laparoscopic videos

Histopathologic inflammation grades were distributed heterogeneously, with intermediate and severe cases predominating in most centers (**Figure 3d**). The internal validation for multiclass histopathological appendicitis grading revealed distinct center- and grade-specific variation in local model performance. Overall, both the centralized and the Swarm Learning models outperformed local models at average AUROCs of [0.784] ± [0.008] and [0.775] ± [0.011], respectively (**Figure 3e, Supplementary Table 6**).

Upon external validation, we observed similar grade-specific performance patterns. Swarm Learning models outperformed local models on average, with particularly reliable classification of Grade 3 (AUROC: [0.731] ± [0.023]) and Grade 0 (AUROC: [0.676] ± [0.047]). While some models trained on data from individual centers showed relatively high average AUROC (e.g., Center 2, [0.654] ± [0.014]), the Swarm Learning and centralized models achieved the highest overall average performances across all grades at [0.626] ± [0.029] and [0.610] ± [0.018], respectively (**Figure 3f, Supplementary Table 6**). These findings underscore the generalizability and robustness of Swarm Learning, as it consistently outperformed or matched local models across multiple inflammation grades.

### Hardware reliability and system integration as key challenges for the clinical adoption of Swarm Learning

Following hardware installation, three out of five participating centers required at least one hardware replacement during the study. The majority of Swarm Learning internet connections relied on Wi-Fi rather than LAN (**Figure 4a**). While both the graphical user interface and hardware components were predominantly rated as reliable, internet connectivity received more variable reliability ratings (**Figure 4b**).

**Figure 4:**
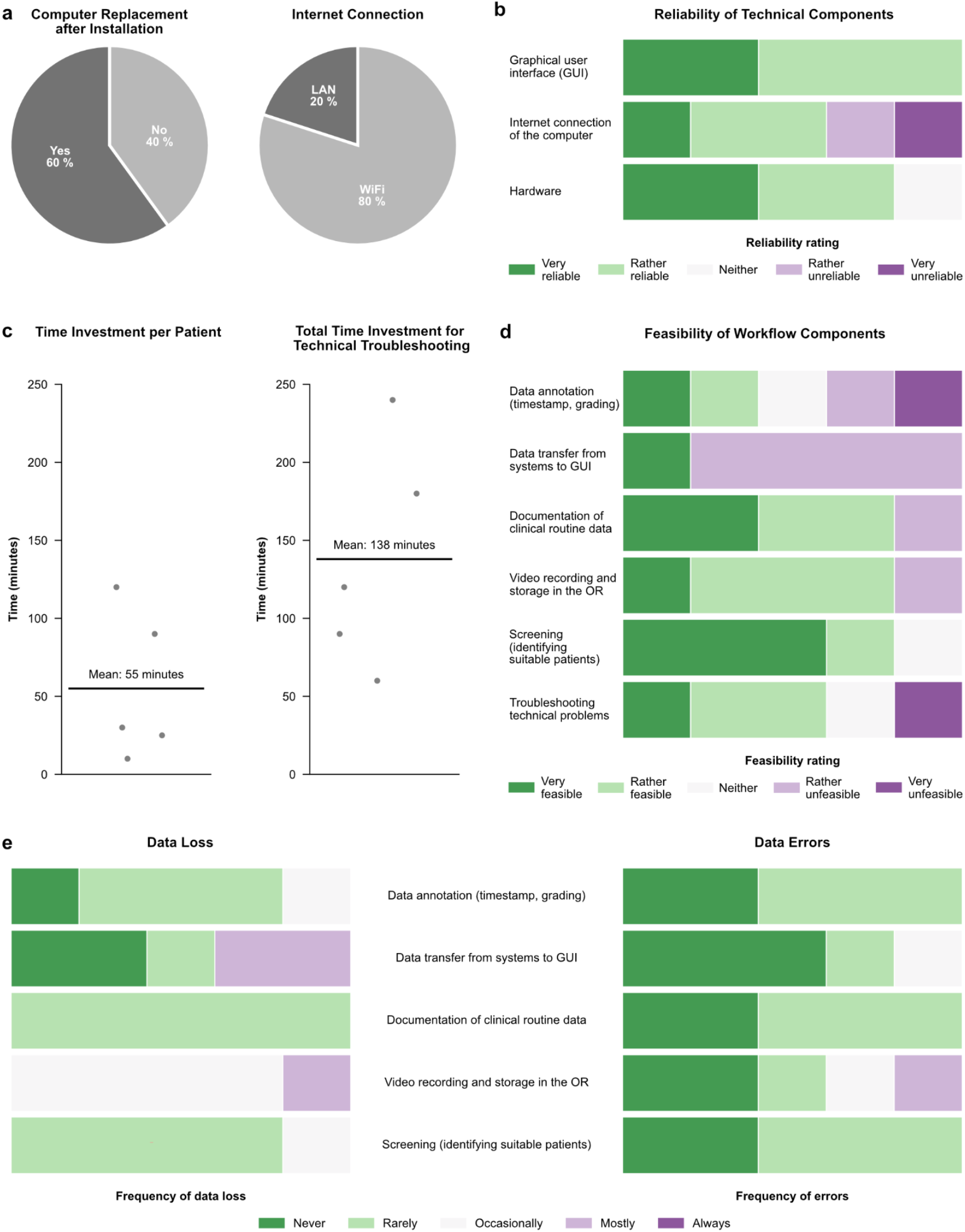
Survey results on technical reliability, feasibility, and workload of the decentralized digital system. **(a)** Pie charts showing the proportion of centers that required computer replacement after installation (left) and the type of internet connection used (Wi-Fi vs. LAN, right). **(b)** User ratings of the reliability of technical components, including the graphical user interface (GUI), computer internet connection, and hardware, on a five-point Likert scale from “very reliable” to “very unreliable”. **(c)** Dot plots of the time investment per patient for study-related tasks (left) and total time investment for technical troubleshooting per center (right); black lines indicate mean values. **(d)** Feasibility ratings for workflow components, including data annotation, data transfer, documentation of routine clinical data, video recording and storage, patient screening, and troubleshooting, on a five-point Likert scale from “very feasible” to “very unfeasible”. **(e)** Reported frequency of data loss (left) and data errors (right) across workflow components, with responses ranging from “never” to “always”.

Participation in the study increased the workload for the contributing centers, with an estimated average of 55 minutes spent on study-related tasks per patient and 138 minutes on troubleshooting technical issues per center during the study period (**Figure 4c**). Several workflow components were not consistently considered feasible, especially those related to video and clinical data documentation, annotation, and transfer from clinical systems to the graphical user interface (**Figure 4d**). The contributing centers identified the most significant risk of data loss in video recording, data annotation, and data transfer from clinical systems to the graphical user interface (**Figure 4e**).

These findings emphasize local infrastructure, including hardware and internet connectivity, as well as limited integration of the study workflow with electronic health records and laparoscopic video infrastructure, as key failure points in data collection and decentralized surgical video analysis.

## Discussion

This international multicenter study represents the first evaluation of Swarm Learning^23^, a decentralized, privacy-preserving deep learning method, for collaborative analysis of laparoscopic video data. Using a novel multicenter dataset, we introduce disease stage prediction from appendectomy videos as a new prediction task and benchmark Swarm Learning against local and centralized learning strategies. Additionally, we identify technical and workflow-related obstacles to the clinical translation of decentralized learning for surgical video analysis through a comprehensive user survey.

Models trained using Swarm Learning consistently matched or outperformed the performance of local and centralized models across patient-level prediction tasks. These findings indicate the potential of Swarm Learning to facilitate multicenter collaboration for surgical video analysis. In this field, liability and data privacy concerns are particularly prevalent because surgery recordings not only inform about the patient but also about the surgical team’s performance^21^.

Detection of perforated appendicitis, laparoscopic grading, and histopathologic inflammation grading are patient-level prediction tasks. Using patient-level labels in a weakly supervised framework drastically reduces the annotation burden for surgical video data compared to frame-level labels. However, most current research in surgical video analysis focuses on frame-level prediction tasks with limited or indirect clinical relevance, such as instrument segmentation or anatomy detection.^16^ Considering the status quo in surgical video analysis research, which is often based on small, single-center datasets, our study represents a major step forward. Our results show that multicenter training, both via centralized and decentralized approaches, improves model generalizability and robustness, particularly in scenarios with limited or imbalanced data.

Swarm Learning offers a promising alternative to traditional federated learning by eliminating the need for a central coordinator, thereby reducing vulnerability to single points of failure and aligning more closely with real-world clinical governance constraints. Automated extraction of patient-level labels from surgery reports or histopathologic images could further reduce the annotation burden on clinicians (surgeons and pathologists). While laparoscopic and histopathologic appendicitis grading may have limited relevance for therapeutic decision-making in the treatment of acute appendicitis, our approach is transferable to other, more complex prediction tasks where temporospatial information from video data (i.e., encoding the technical quality of an operation) contributes to objectifiable and relevant patient outcomes. Prior studies have shown that surgeon skill, as captured in video, correlates with postoperative outcomes in esophageal^28^, bariatric^29^, and colorectal surgery^30^. In light of this evidence, our approach could be extended to support quality assurance and outcome prediction in minimally invasive surgery.

We observed considerable heterogeneity in model performance across centers and disease stages, particularly for histopathologic grading. This variability likely reflects differences in patient populations, surgical practices, and annotation consistency. For example, laparoscopic staging is inherently subjective, as evidenced by moderate inter-rater agreement (weighted Cohen’s kappa of 0.62) in the Appendix300 dataset^27^. Moreover, the lower performance of all models on histopathologic grading compared to laparoscopic grading suggests that predicting pathologic properties reflecting biological processes from macroscopic video features remains a challenging and indirect task. This observation supports the notion that inflammation processes during acute appendicitis are neither linear processes nor all (microscopic) pathologic processes manifest in a macroscopic, laparoscopically visible phenotype^31,32^.

Several practical challenges and limitations remain. Our survey revealed that hardware reliability, internet connectivity, and lack of integration with clinical systems were major barriers to implementation. The average troubleshooting time exceeded two hours per center. Evidently, decentralized learning pipelines for surgical AI need to integrate with electronic health records and surgical video recording infrastructure and avoid local hardware dependencies, potentially via cloud-based solutions. As a proof-of-concept, our study is limited in scope. Incorporating outcomes of immediate clinical relevance, such as complications or readmissions, could increase the direct impact of our findings on patient care.

In conclusion, our study establishes technical foundations for privacy-preserving, collaborative AI development in surgical video analysis. By demonstrating the feasibility and benefits of Swarm Learning for weakly supervised, patient-level prediction tasks, we provide a pathway toward more generalizable, robust, and clinically valuable AI tools in surgery. Continued multicenter collaboration and methodological innovation will be critical to translating these advances into routine clinical practice and ultimately improve surgical outcomes.

## Data Availability

A detailed data descriptor outlining the recording, annotation, and technical validation process is available at https://doi.org/10.1101/2025.09.05.25335174. The development cohort multicenter dataset of appendectomy recordings and corresponding clinical metadata will be made publicly available upon acceptance of this work.

## Acknowledgements

MK and ACJ are supported by the European Union through NEARDATA under grant agreement ID 101092644. JNK is supported by the German Cancer Aid DKH (DECADE, 70115166), the German Federal Ministry of Research, Technology and Space BMFTR (PEARL, 01KD2104C; CAMINO, 01EO2101; TRANSFORM LIVER, 031L0312A; TANGERINE, 01KT2302 through ERANET Transcan; Come2Data, 16DKZ2044A; DEEP-HCC, 031L0315A; DECIPHER-M, 01KD2420A; NextBIG, 01ZU2402A), the German Research Foundation DFG (CRC/TR 412, 535081457; SFB 1709/1 2025, 533056198), the German Academic Exchange Service DAAD (SECAI, 57616814), the German Federal Joint Committee G-BA (TransplantKI, 01VSF21048), the European Union EU’s Horizon Europe research and innovation programme (ODELIA, 101057091; GENIAL, 101096312), the European Research Council ERC (NADIR, 101114631), the National Institutes of Health NIH (EPICO, R01 CA263318) and the National Institute for Health and Care Research NIHR (Leeds Biomedical Research Centre, NIHR203331). FRK receives support from the German Cancer Research Center (CoBot 2.0), the Joachim Herz Foundation (Add-On Fellowship for Interdisciplinary Life Science), the Central Indiana Corporate Partnership AnalytiXIN Initiative, the Evan and Sue Ann Werling Pancreatic Cancer Research Fund, and the Indiana Clinical and Translational Sciences Institute (EPAR4157) funded, in part, by Grant Number UM1TR004402 from the National Institutes of Health, National Center for Advancing Translational Sciences, Clinical and Translational Sciences Award. The views expressed are those of the author(s) and not necessarily those of the National Institutes of Health, the NHS, the NIHR, or the Department of Health and Social Care. This work was funded by the European Union. Views and opinions expressed are, however, those of the author(s) only and do not necessarily reflect those of the European Union. Neither the European Union nor the granting authority can be held responsible for them.

