## Supplementary Material for "Decentralized, privacy-preserving surgical video analysis with Swarm Learning"

### **Supplementary Information 1:** Patient cohort and data preprocessing

#### Patient cohort

A total of 392 patients with suspected acute appendicitis undergoing laparoscopic appendectomy at six independent clinical centers were recruited. In addition, five patients undergoing non-appendectomy laparoscopic surgery were recruited as negative controls, resulting in a total of 397 laparoscopic surgery recordings analyzed in this study. Five German centers provided data for the development cohort used for model implementation and internal validation (Asklepios-ASB Krankenhaus Radeberg; Diakonissenkrankenhaus Dresden; Krankenhaus St. Joseph Stift Dresden; St. Elisabethen-Krankenhaus Ravensburg; Department of Pediatric Surgery, University Hospital Carl Gustav Carus Dresden). One Portuguese center (Hospital Prof. Dr. Fernando Fonseca, Lisbon) provided data for external testing. All included patients had a clinical indication for the surgical procedure.

A detailed data descriptor outlining the recording, preprocessing, annotation, and technical validation process is available at https://doi.org/10.1101/2025.09.05.25335174[1]. In brief, appendectomy recordings and clinical metadata, including histopathologic inflammation stage and laparoscopic grade of appendicitis, were consecutively gathered in clinical routine. Patients with an indication of laparoscopic appendectomy for suspected acute appendicitis and documented histopathologic inflammation grade were included in the appendectomy dataset. Exclusion criteria comprised conversion to open surgery, incomplete surgery recordings, and cases with corrupted files.

Surgeries were performed, recorded, and saved using locally available laparoscopic hardware. All data were compiled in an anonymized fashion using a graphical user interface. Through this graphical user interface, a timestamp at which the appendix is fully visible prior to invasive preparation, was documented. Based on this timestamp, a 100-second video snippet was extracted from the full video recording, which covers 50 seconds before and after the time point of full appendix visibility. Laparoscopic appendicitis grades were labeled by two independent annotators (general surgery residents or fellows with at least 4 years of experience in laparoscopic surgery) based on the appendectomy recording. Disagreements were resolved through a third annotation by a general surgery resident with 7 years of experience in laparoscopic surgery. Histopathologic inflammation grades were derived from routine pathology reports.

#### Preprocessing workflow

To protect patient and caregiver privacy, videos were anonymized using a publicly available out-of-body detector[2], which automatically masked areas outside the operative field. Additionally, any laparoscopic system interface overlays containing patient-identifiable information were manually cropped before processing. Subsequently, 200 video frames were extracted from the 100-second video snippet centered around the annotated timestamp using Fast Forward Moving Picture Experts Group (FFmpeg), an open-source multimedia framework. This frame window provided relevant surgical context while keeping data size manageable.

All patient metadata were merged into a unified tabular format to support efficient data handling and facilitate the division of samples into training, validation, and test sets for machine learning model development.

### **Supplementary Information 2:** Benchmarking of temporal resolution and modeling strategies for the detection of perforated appendicitis

To determine optimal experimental parameters for all subsequent experiments, we systematically evaluated the impact of temporal information and frame sampling strategies on binary surgical video classification performance by investigating six different frame sampling configurations (1, 10, 25, 50, 100, and 200 frames, corresponding to frame rates of 0.1-2 frames per second) on the binary prediction task (detection of perforated appendicitis). The middle frame was always included, and additional frames were sampled symmetrically around it. Across these frame sampling configurations, we benchmarked three AI methods, as detailed in **Supplementary Information 3** (Multiple Instance Learning, SurgFrameNet, SurgTempoNet).

These configuration experiments were validated on a combined internal validation dataset comprising 20% of the data from each center in the development cohort, selected randomly. Each selected model was trained three times using 80% of the data from each center from the development cohort (five German centers), with internal validation performed on the remaining 20%. The training cohort included a total of 261 patients, while the internal validation cohort comprised 69 patients (**Figure 1a**).

Using the best-performing configuration, we conducted leave-one-center-out cross-validation experiments to evaluate the cross-institutional generalizability of patient-level prediction models for the detection of perforated appendicitis, where each center was held out once for validation while models were trained on data from the remaining centers in the development cohort.

**Supplementary Tables 1-3** present the area under the receiver operating characteristic curve (AUROC) values for each technique across all frame rates. Based on these results, the SurgTempoNet models consistently outperformed their SurgFrameNet and MIL counterparts (**Figure 2a**, **Supplementary Tables 1-3**). Among all evaluated models, the SurgTempoNet configuration using 1.0 fps achieved the highest AUROC of [0.918] ± [0.007]. The other two approaches, SurgFrameNet and MIL, yielded lower performances of [0.905] ± [0.006] and [0.800] ± [0.035], respectively, at a frame rate of 1.0 fps (**Supplementary Tables 1-3**).

These results indicate that frame sampling at 1.0 fps results in reliable classification performance for the detection of perforated appendicitis, with SurgTempoNet consistently achieving superior results compared to SurgFrameNet and MIL across all centers (**Figure 2a**, **Supplementary Table 2**). Therefore, this configuration was selected for all subsequent experiments.

### **Supplementary Information 3:** AI methods

**Multiple instance learning (MIL) method:** MIL[3,4] has emerged as a promising supervised learning framework for developing deep learning algorithms in medical applications, particularly when precise frame-level annotations are ambiguous or unavailable. Unlike conventional supervised learning, which requires a distinct class label for each individual observation or "instance", MIL assigns labels to collections of observations, referred to as "bags of instances"[31], where the label reflects the collective properties of the group rather than any single instance within it. In our approach, MIL operates in two phases: feature extraction and training. First, video frames are transformed into 512-dimensional feature vectors using a ResNet-18 model pre-trained on ImageNet[5]. These extracted features are then used to train a MIL model to predict outcomes at the patient level. The model employs an attention-based MIL architecture (Att-MIL)[6], consisting of a multilayer perceptron classifier network (512×256, followed by 256×2) and an attention mechanism[7] to weigh the significance of each instance within a bag. This process involves aggregating predictions from multiple frames belonging to the same video, resulting in a comprehensive, patient-level prediction for each surgical case. By leveraging the MIL framework, our method addresses the challenges of weak supervision and enhances the robustness and interpretability of outcome prediction in medical video analysis.

**SurgNet method:** This study introduces the SurgNet framework for surgical video analysis, comprising two deep learning techniques: SurgTempoNet, which models both spatial and temporal patterns, and SurgFrameNet, which focuses on spatial features alone. Both approaches are built upon a hybrid PhaseLSTMConvNext architecture that integrates the spatial feature extraction power of convolutional networks (ConvNet) with the temporal modeling capabilities of Long Short-Term Memory (LSTM) networks[8,9]. The ConvNet backbone, pre-trained on ImageNet-22k and fine-tuned on ImageNet-1k at a resolution of 384×384 pixels, serves as the feature extractor, with early layers frozen to retain foundational representations. **SurgTempoNet** leverages temporal modeling by passing extracted 1024-dimensional spatial features through an LSTM layer with 160 hidden units, enabling the network to capture sequential dependencies and temporal patterns across video frames. In contrast, **SurgFrameNet** is a non-temporal variant that bypasses the LSTM, feeding spatial features directly to the classification head for frame-wise, independent analysis. Both models were trained using the AdamW optimizer (learning rate 0.001, batch size 64, 64 epochs)[10] and employed class-weighted cross-entropy loss to address data imbalance. Data preprocessing included standard ImageNet normalization, augmentation, and resizing frames to 480×270 pixels for computational efficiency. Model selection was based on area under the receiver operating curve (AUROC)-score performance on validation sets, with the best-performing models retained for final evaluation.

**Supplementary Information 4:** Swarm Learning workflow

In this study, we investigate the potential of Swarm Learning in co-training machine learning models for the purpose of predicting patient-level outcomes (disease stages) from surgical video data (appendectomy recordings). We implemented a Swarm Learning network consisting of five separate "nodes", and trained a model using this network (**Figure 1e**). During the training process, model weights were exchanged between nodes at multiple synchronization events (sync events), which took place at the end of each synchronization interval. The synchronization interval represents the number of batches after which learning sharing occurs. The model weights were then averaged at each sync event, and training continued at each node using the averaged parameters. We utilized a weighted averaging approach, which involved multiplying each node's weights by a weighting factor proportional to the amount contributed by the partner. This approach was motivated by previous studies in breast, gastric, and colorectal cancer pathology[11–13].

Our Swarm Learning implementation stored metadata about the model synchronization on an Ethereum blockchain, with the blockchain managing the global status information about the model. We used the Hewlett Packard Enterprise (HPE) Swarm Learning implementation, which consisted of four components: the Swarm Learning process, the Swarm Network (SN) process, identity management, and HPE license management. All processes, or nodes, were run in multiple Docker containers. We provide a detailed description of our Swarm Learning process, along with a small sample dataset and instructions on how to reproduce our experiments using our code.

**Supplementary Table 1:** SurgFrameNet

| Frame sampling rate (fps) | Training Center | | | | | | | | | | | |
|---|---|---|---|---|---|---|---|---|---|---|---|---|
| | Centralized | | Center 1 | | Center 2 | | Center 3 | | Center 4 | | Center 5 | |
| | mean | std | mean | std | mean | std | mean | std | mean | std | mean | std |
| **Center frame** | 0.830 | 0.019 | 0.661 | 0.152 | 0.783 | 0.003 | 0.675 | 0.076 | 0.787 | 0.057 | 0.707 | 0.059 |
| **0.1** | 0.896 | 0.009 | 0.81 | 0.011 | 0.698 | 0.095 | 0.818 | 0.055 | 0.868 | 0.005 | 0.852 | 0.023 |
| **0.25** | 0.903 | 0.007 | 0.837 | 0.027 | 0.707 | 0.051 | 0.809 | 0.034 | 0.85 | 0.012 | 0.792 | 0.079 |
| **0.5** | 0.899 | 0.019 | 0.81 | 0.025 | 0.795 | 0.076 | 0.782 | 0.049 | 0.86 | 0.003 | 0.825 | 0.039 |
| **1.0** | 0.905 | 0.006 | 0.812 | 0.004 | 0.505 | 0.193 | 0.791 | 0.02 | 0.851 | 0.005 | 0.857 | 0.005 |
| **2.0** | 0.911 | 0.006 | 0.814 | 0.007 | 0.788 | 0.074 | 0.768 | 0.053 | 0.855 | 0.007 | 0.833 | 0.026 |

**Supplementary Table 1: Benchmarking of temporal sampling strategies for detection of perforated appendicitis from appendectomy recordings with SurgFrameNet models.** Table displays mean AUROC values and standard deviations for SurgFrameNet models trained on pooled training data subsets from five centers and tested on 20% of data from all centers for each temporal sampling strategy.

### **Supplementary Table 2:** SurgTempoNet

| **Frame sampling rate (fps)** | **Training Center** | | | | | | | | | | | |
|---|---|---|---|---|---|---|---|---|---|---|---|---|
| | Centralized | | Center 1 | | Center 2 | | Center 3 | | Center 4 | | Center 5 | |
| | mean | std | mean | std | mean | std | mean | std | mean | std | mean | std |
| **Center frame** | 0.868 | 0.008 | 0.829 | 0.02 | 0.796 | 0.049 | 0.864 | 0.004 | 0.815 | 0.02 | 0.799 | 0.025 |
| **0.1** | 0.903 | 0.011 | 0.818 | 0.008 | 0.839 | 0.035 | 0.891 | 0.007 | 0.868 | 0.009 | 0.864 | 0.084 |
| **0.25** | 0.904 | 0.005 | 0.828 | 0.003 | 0.831 | 0.038 | 0.892 | 0.001 | 0.867 | 0.006 | 0.842 | 0.037 |
| **0.5** | 0.917 | 0.007 | 0.823 | 0.014 | 0.861 | 0.033 | 0.888 | 0.007 | 0.878 | 0.006 | 0.877 | 0.025 |
| **1.0** | 0.918 | 0.007 | 0.828 | 0.014 | 0.794 | 0.133 | 0.89 | 0.004 | 0.879 | 0.001 | 0.804 | 0.05 |
| **2.0** | 0.901 | 0.018 | 0.816 | 0.005 | 0.857 | 0.002 | 0.869 | 0.012 | 0.876 | 0.015 | 0.879 | 0.042 |

**Supplementary Table 2: Benchmarking of temporal sampling strategies for detection of perforated appendicitis from appendectomy recordings with SurgTempoNet models.** Table displays mean AUROC values and standard deviations for SurgTempoNet models trained on pooled training data subsets from five centers and tested on 20% of data from all centers for each temporal sampling strategy.

### **Supplementary Table 3:** Multiple Instance Learning

| Frame sampling rate (fps) | Training Center | | | | | | | | | | | |
|---|---|---|---|---|---|---|---|---|---|---|---|---|
| | Centralized | | Center 1 | | Center 2 | | Center 3 | | Center 4 | | Center 5 | |
| | mean | std | mean | std | mean | std | mean | std | mean | std | mean | std |
| **Center frame** | 0.712 | 0.035 | 0.537 | 0.106 | 0.642 | 0.062 | 0.542 | 0.11 | 0.551 | 0.087 | 0.593 | 0.033 |
| **0.1** | 0.801 | 0.034 | 0.564 | 0.032 | 0.744 | 0.059 | 0.758 | 0.032 | 0.75 | 0.029 | 0.745 | 0.119 |
| **0.25** | 0.791 | 0.02 | 0.521 | 0.182 | 0.694 | 0.064 | 0.771 | 0.053 | 0.777 | 0.015 | 0.73 | 0.085 |
| **0.5** | 0.795 | 0.026 | 0.557 | 0.098 | 0.767 | 0.042 | 0.751 | 0.008 | 0.77 | 0.008 | 0.776 | 0.027 |
| **1.0** | 0.8 | 0.035 | 0.621 | 0.055 | 0.714 | 0.051 | 0.756 | 0.049 | 0.768 | 0.025 | 0.748 | 0.046 |
| **2.0** | 0.795 | 0.037 | 0.588 | 0.108 | 0.683 | 0.096 | 0.758 | 0.04 | 0.75 | 0.021 | 0.753 | 0.031 |

**Supplementary Table 3: Benchmarking of sampling strategies for detection of perforated appendicitis from appendectomy recordings with Multiple Instance Learning (MIL) models.** Table displays mean AUROC values and standard deviations for MIL models trained on pooled training data subsets from five centers and tested on 20% of data from all centers for each temporal sampling strategy.

**Supplementary Table 4:** Comparison of local, centralized, and Swarm Learning for detection of perforated appendicitis

| Training Center | F1-score | | AUROC | | p-value |
|---|---|---|---|---|---|
| | mean | std | mean | std | Two-sided t-test for AUROC comparison with the Swarm Learning model |
| **Center 1** | 0.4751 | 0.0063 | 0.9529 | 0.0098 | 0.163607 |
| **Center 2** | 0.6963 | 0.1876 | 0.9713 | 0.0063 | 0.416328 |
| **Center 3** | 0.6353 | 0.1016 | 0.9483 | 0.0384 | 0.517764 |
| **Center 4** | 0.6996 | 0.0915 | 0.9428 | 0.0500 | 0.512879 |
| **Center 5** | 0.5542 | 0.0200 | 0.8504 | 0.0829 | 0.136216 |
| **Centralized** | 0.5956 | 0.0467 | 0.9443 | 0.0035 | 0.034132 |
| **Swarm Learning** | 0.6360 | 0.1350 | 0.9657 | 0.0085 | 1 |

**Supplementary Table 4: Performance metrics for detection of perforated appendicitis from appendectomy recordings and statistical comparison of models by training center.** This table presents key performance metrics, including the F1-score and AUROC (both reported as mean ± standard deviation), for models trained at individual Training Centers (Center 1 to Center 5), an aggregate centralized model, and a Swarm Learning model baseline. Additionally, the table includes the p-value derived from a two-sided t-test conducted for each center's AUROC score in comparison to the swarm model's AUROC, providing a statistical assessment of differences in performance.

**Supplementary Table 5:** Comparison of local, centralized, and Swarm Learning for laparoscopic appendicitis grading

| Center | Internal validation AUROC | | | | | | |
|---|---|---|---|---|---|---|---|
| | Grade 0 | Grade 1 | Grade 2 | Grade 3 | Grade 4 | Grade 5 | Average |
| Center 1 | 0.717 ± 0.002 | 0.777 ± 0.018 | 0.623 ± 0.009 | 0.607 ± 0.014 | 0.574 ± 0.084 | 0.513 ± 0.091 | 0.635 ± 0.023 |
| Center 2 | 0.664 ± 0.018 | 0.756 ± 0.046 | 0.438 ± 0.016 | 0.634 ± 0.034 | 0.803 ± 0.027 | 0.522 ± 0.139 | 0.636 ± 0.030 |
| Center 3 | 0.490 ± 0.055 | 0.745 ± 0.024 | 0.582 ± 0.096 | 0.621 ± 0.021 | 0.841 ± 0.010 | 0.370 ± 0.125 | 0.608 ± 0.048 |
| Center 4 | 0.478 ± 0.056 | 0.704 ± 0.039 | 0.608 ± 0.057 | 0.560 ± 0.075 | 0.764 ± 0.056 | 0.663 ± 0.043 | 0.629 ± 0.050 |
| Center 5 | 0.561 ± 0.019 | 0.825 ± 0.025 | 0.614 ± 0.031 | 0.635 ± 0.041 | 0.826 ± 0.010 | 0.475 ± 0.086 | 0.656 ± 0.030 |
| Centralized | 0.717 ± 0.000 | 0.879 ± 0.000 | 0.670 ± 0.002 | 0.672 ± 0.048 | 0.864 ± 0.012 | 0.732 ± 0.049 | 0.756 ± 0.017 |
| Swarm Learning | 0.687 ± 0.009 | 0.878 ± 0.006 | 0.648 ± 0.038 | 0.684 ± 0.024 | 0.809 ± 0.016 | 0.678 ± 0.002 | 0.731 ± 0.013 |

| Center | External validation AUROC | | | | | | |
|---|---|---|---|---|---|---|---|
| | Grade 0 | Grade 1 | Grade 2 | Grade 3 | Grade 4 | Grade 5 | Average |
| Center 1 | NA | 0.698 ± 0.029 | 0.718 ± 0.028 | 0.776 ± 0.109 | 0.182 ± 0.226 | 0.677 ± 0.042 | 0.610 ± 0.210 |
| Center 2 | NA | 0.345 ± 0.041 | 0.667 ± 0.013 | 0.804 ± 0.020 | 0.859 ± 0.027 | 0.699 ± 0.020 | 0.675 ± 0.175 |
| Center 3 | NA | 0.678 ± 0.026 | 0.546 ± 0.020 | 0.853 ± 0.015 | 0.101 ± 0.072 | 0.607 ± 0.041 | 0.557 ± 0.250 |
| Center 4 | NA | 0.576 ± 0.014 | 0.449 ± 0.015 | 0.879 ± 0.014 | 0.813 ± 0.011 | 0.693 ± 0.010 | 0.682 ± 0.165 |
| Center 5 | NA | 0.731 ± 0.023 | 0.713 ± 0.010 | 0.863 ± 0.013 | 0.232 ± 0.304 | 0.681 ± 0.046 | 0.644 ± 0.225 |
| Centralized | NA | 0.732 ± 0.008 | 0.687 ± 0.061 | 0.918 ± 0.012 | 0.833 ± 0.014 | 0.803 ± 0.067 | 0.795 ± 0.092 |
| Swarm Learning | NA | 0.701 ± 0.015 | 0.746 ± 0.008 | 0.892 ± 0.020 | 0.924 ± 0.033 | 0.825 ± 0.017 | 0.818 ± 0.092 |

**Supplementary Table 5: Internal and external validation AUROC scores for laparoscopic grading across local learning, centralized learning, and Swarm Learning approaches.** This table presents the mean AUROC scores (± standard deviation) for laparoscopic grading, evaluated across different centers (Center 1-Center 5), a centralized model, and Swarm Learning. The top section of the table details the internal validation AUROC scores, providing performance metrics for each laparoscopic grade (Grade 0-5) and an overall average AUROC score across these grades. Correspondingly, the bottom section reports the external validation AUROC scores, displaying the mean AUROC (± standard deviation) for each laparoscopic grade and the average performance specifically within the external validation cohort.

**Supplementary Table 6:** Comparison of local, centralized, and Swarm Learning for histopathologic inflammation grading

| Center | Internal validation AUROC | | | | |
|---|---|---|---|---|---|
| | Grade 0 | Grade 1 | Grade 2 | Grade 3 | Average |
| **Center 1** | 0.565 ± 0.111 | 0.830 ± 0.021 | 0.559 ± 0.049 | 0.662 ± 0.055 | 0.654 ± 0.041 |
| **Center 2** | 0.633 ± 0.082 | 0.780 ± 0.015 | 0.595 ± 0.010 | 0.702 ± 0.018 | 0.677 ± 0.018 |
| **Center 3** | 0.336 ± 0.055 | 0.865 ± 0.006 | 0.521 ± 0.020 | 0.674 ± 0.027 | 0.599 ± 0.027 |
| **Center 4** | 0.577 ± 0.040 | 0.704 ± 0.015 | 0.494 ± 0.008 | 0.724 ± 0.015 | 0.625 ± 0.021 |
| **Center 5** | 0.503 ± 0.064 | 0.773 ± 0.038 | 0.470 ± 0.038 | 0.680 ± 0.020 | 0.606 ± 0.003 |
| **Centralized** | 0.949 ± 0.015 | 0.763 ± 0.018 | 0.656 ± 0.041 | 0.768 ± 0.020 | 0.784 ± 0.008 |
| **Swarm Learning** | 0.884 ± 0.090 | 0.810 ± 0.050 | 0.633 ± 0.036 | 0.772 ± 0.027 | 0.775 ± 0.011 |

| Center | External validation AUROC | | | | |
|---|---|---|---|---|---|
| | Grade 0 | Grade 1 | Grade 2 | Grade 3 | Average |
| **Center 1** | 0.620 ± 0.093 | 0.599 ± 0.027 | 0.483 ± 0.007 | 0.686 ± 0.032 | 0.597 ± 0.038 |
| **Center 2** | 0.730 ± 0.024 | 0.635 ± 0.008 | 0.550 ± 0.016 | 0.700 ± 0.009 | 0.654 ± 0.014 |
| **Center 3** | 0.599 ± 0.104 | 0.629 ± 0.006 | 0.501 ± 0.016 | 0.736 ± 0.020 | 0.616 ± 0.037 |
| **Center 4** | 0.503 ± 0.038 | 0.631 ± 0.006 | 0.523 ± 0.009 | 0.706 ± 0.004 | 0.591 ± 0.014 |
| **Center 5** | 0.544 ± 0.107 | 0.562 ± 0.017 | 0.502 ± 0.033 | 0.695 ± 0.020 | 0.576 ± 0.044 |
| **Centralized** | 0.661 ± 0.012 | 0.565 ± 0.014 | 0.508 ± 0.009 | 0.707 ± 0.036 | 0.610 ± 0.018 |
| **Swarm Learning** | 0.676 ± 0.047 | 0.545 ± 0.027 | 0.551 ± 0.019 | 0.731 ± 0.023 | 0.626 ± 0.029 |

**Supplementary Table 6: Internal and external validation AUROC scores for histopathologic inflammation grading across local learning, centralized learning, and Swarm Learning approaches.** This table presents the mean AUROC scores (± standard deviation) for multiclass histopathological grading, evaluated across different centers (Center 1-Center 5), a centralized model, and Swarm Learning. The top section of the table details the internal validation AUROC scores, providing performance metrics for each histopathologic inflammation grade (Grade 0-3) and an overall average AUROC score across these grades. Correspondingly, the bottom section reports the external validation AUROC scores, displaying the mean AUROC (± standard deviation) for each histopathologic inflammation grade and the average performance specifically within the external validation cohort.
